# Updated outcome of patients with lung only osteosarcoma recurrence following surgical resection enrolled in phase 2 clinical trials: A report from the Children’s Oncology Group

**DOI:** 10.64898/2026.11.13.26368045

**Authors:** Alexander J. Chou, Natalie DelRocco, Mark Krailo, Allen Buxton, Pooja Hingorani, Brenda J. Weigel, Elizabeth Fox, Joanne P Lagmay, Richard Gorlick, Ryan Roberts, Steven G. DuBois, Damon R. Reed, Katherine Janeway

**Affiliations:** Weill Cornell Medicine; Department of Population and Public Health Sciences, University of Southern California, Los Angeles, CA; Children’s Oncology Group; AbbVie Inc., North Chicago, IL; Department of Oncology, St. Jude Children’s Research Hospital, Memphis, TN; UF Health Shands Children’s Hospital, Gainesville, FL; MD Anderson Cancer Center; Nationwide Children’s Hospital and The Ohio State University, Columbus, OH; Dana-Farber / Boston Children’s Cancer and Blood Disorders Center and Harvard Medical School; Memorial Sloan Kettering Cancer Center, Department of Pediatrics, New York, NY, 10065

**Keywords:** benchmarks, recurrent osteosarcoma, Children’s Oncology Group, relapse, adjuvant, trial design

## Abstract

**Objective:** Osteosarcoma is a rare primary bone cancer of adolescents and young adults without significant improvement in treatments or outcomes in several decades. Resection is standard of care for recurrent disease. Patients with lung only osteosarcoma recurrence following surgical resection (LOOR-FSR) may present the most meaningful opportunity for impactful intervention but alternative trial endpoints to objective response are needed. Here we update benchmarks for LOOR-FSR with contemporary clinical trial data.

**Patients/Methods:** This study included patients with pathologically confirmed LOOR-FSR enrolled on three Children’s Oncology Group phase 2 single arm trials. Benchmark endpoints assessed in this patient population were event-free survival (EFS) and overall survival (OS). In addition, we evaluated the impact of demographics and baseline clinical factors on outcome.

**Results:** Ninety-nine patients met criteria for inclusion in this analysis. Median EFS and OS were 4.8 (95% CI: 3.5, 6.3) and 26.2 (23.3, 56.8) months respectively. Twelve-month EFS was 22% (15-31%) and ranged from 14% to 29% across the 3 trials, a difference that did not achieve statistical significance. The risk of EFS event was higher for those whose first recurrence occurred within two years of diagnosis (p=0.009). Other demographic and clinical factors did not statistically impact risk of event though estimated EFS for males was lower than for females.

**Conclusion:** Despite almost doubling the number of patients in this historical control cohort, wide variance in outcomes led to confidence intervals unchanged from previously published work. Until the most significant factors contributing to variability of outcomes in this patient population are quantified, single arm trials of new approaches will only be able to identify large effect sizes. Smaller effect sizes will require randomization.

## INTRODUCTION

Thirty to forty percent of patients with high-grade osteosarcoma will eventually experience disease recurrence.^1^ The majority of these patients will relapse with lung-only disease^1–5^. For many patients, complete surgical resection of pulmonary sites of recurrent metastatic disease is possible and considered standard of care.^6^ Patients with lung-only osteosarcoma recurrence following surgical resection (LOOR-FSR) represent a population with more favorable outcomes compared with patients with widespread and/or unresectable relapse^4,5^. Moreover, patients with LOOR-FSR may be biologically similar to patients with newly diagnosed localized osteosarcoma, 80% of whom have microscopic pulmonary metastatic disease at the time of diagnosis. Treatments targeting the biology of microscopic and emerging pulmonary metastasis are identified as high priority for osteosarcoma drug development.^7^ As these drugs may be less active in bulky disease, clinical trials in LOOR-FSR are required to assess their activity.

Identifying novel effective agents in osteosarcoma has been challenging.^2,7,8^ One reason for this is the lack of appropriate clinical trial endpoints for this rare disease. Conventional RECIST criteria are poor surrogates for therapeutic efficacy in osteosarcoma^9^ that is not resected due to its bone matrix. The use of objective response is further complicated patients with LOOR-FSR who do not have no evidence of disease. To facilitate the design of signal finding clinical trials in recurrent/refractory osteosarcoma, we published in 2016 in the *Journal of Clinical Oncology* a disease control benchmark for patients with LOOR-FSR receiving ineffective therapy based on a single cooperative group early phase trial (legacy trial, AOST0221).^10,11^

The Children’s Oncology Group (COG) utilized this benchmark to design and conduct 2 prospective single arm phase 2 trials in this patient population: AOST1321 and AOST1421. This standardization of a relapsed trial endpoint has additionally been used in trials outside of COG. Relapsed osteosarcoma remains an unmet need and trial accrual has exceeded forecasts regularly.^12^

In this report, we present an updated disease control benchmark by analyzing data from the 2 contemporary trials, AOST1321 and AOST1421 with data from the legacy trial. In addition, we additionally provide event-free survival (EFS) and overall survival (OS) and assess the association between clinical factors and outcomes in this patient population.

## MATERIALS and METHODS

### Patients

Table 1 summarizes the trials included in this analysis. Methods and results from all trials are previously published.^11,13,14^ AOST0221 (legacy trial) was a phase 2 study that enrolled patients with resectable recurrent osteosarcoma limited to lungs only in first relapse^11^. One of the primary study aims was to determine the effect of inhaled granulocyte macrophage colony stimulating factor (GM-CSF) on EFS. Surgical resection was performed on study after 2 cycles of therapy (6 weeks). The study enrolled patients from 2004 – 2008. AOST1321 was a phase 2 study of denosumab (a monoclonal antibody against receptor activator of nuclear factor-κB ligand (RANKL)) in recurrent/refractory osteosarcoma that enrolled patients from 2015 through 2018. AOST1321 Cohort 2 enrolled patients with fully resected recurrent OS. AOST1421 was a phase 2 study of dinutuximab, a monoclonal antibody against GD2, in combination with GM-CSF in patients with LOOR-FSR. This study enrolled patients from 2015 through 2018, concurrent with AOST1321 with priority for enrollments going to AOST1421 when both were open. Surgical resection was required prior to enrollment on both contemporary studies. Study results have been reported elsewhere. Participants or the participants guardian provided informed consent to the clinical trial protocol and each trial was approval by a local Human Investigations Committee and, when applicable, the Department of Health and Human Services.

**Table 1:** Patient characteristics across trials.

|  | AOST0221<br>(N=37) | AOST1321<br>(N=24) | AOST1421<br>(N=38) | Total<br>(N=99) | P-value |
|---|---|---|---|---|---|
| **Key Trial eligibility** |  |  |  |  |  |
| Recurrence | First | Any | Any |  |  |
| Sites of disease | Lung only | Any | Lung only |  |  |
| **Surveillance intervals** |  |  |  |  |  |
|  | Every 8 wks | Every 8 wks | 6, 15 wks then every 12 wks |  |  |
| **Enrolled Patients (N)** |  |  |  |  |  |
|  | 49 | 38 | 41 | 128 |  |
| **Patients Eligible for analysis (N)** |  |  |  |  |  |
|  | 37 | 24 | 38 | 99 |  |
| **Age in Years at Enrollment**, n (%) |  |  |  |  |  |
| <=9 | 2 (5.4%) | 0 (0.0%) | 4 (10.5%) | 6 (6.1%) | 0.4412<sup>1</sup> |
| 10-17 | 23 (62.2%) | 13 (54.2%) | 21 (55.3%) | 57 (57.6%) |  |
| >=18 | 12 (32.4%) | 11 (45.8%) | 13 (34.2%) | 36 (36.4%) |  |
| **Sex**, n (%) |  |  |  |  |  |
| Male | 25 (67.6%) | 14 (58.3%) | 21 (55.3%) | 60 (60.6%) | 0.5333<sup>1</sup> |
| Female | 12 (32.4%) | 10 (41.7%) | 17 (44.7%) | 39 (39.4%) |  |
| **Race**, n (%) |  |  |  |  |  |
| White | 28 (80.0%) | 18 (94.7%) | 27 (79.4%) | 73 (83.0%) | 0.3704<sup>1</sup> |
| Black | 6 (17.1%) | 1 (5.3%) | 4 (11.8%) | 11 (12.5%) |  |
| Other | 1 (2.9%) | 0 (0.0%) | 3 (8.8%) | 4 (4.5%) |  |
| Missing | 2 | 5 | 4 | 11 |  |
| **Ethnicity**, n (%) |  |  |  |  |  |
| Hispanic or Latino | 2 (5.7%) | 4 (17.4%) | 9 (26.5%) | 15 (16.3%) | 0.0648<sup>1</sup> |
| Not Hispanic or Latino | 33 (94.3%) | 19 (82.6%) | 25 (73.5%) | 77 (83.7%) |  |
| Missing | 2 | 1 | 4 | 7 |  |
| **Metastatic at initial diagnosis**, n (%) |  |  |  |  |  |
| No | 29 (78.4%) | 17 (70.8%) | 29 (76.3%) | 75 (75.8%) | 0.7939<sup>1</sup> |
| Yes | 8 (21.6%) | 7 (29.2%) | 9 (23.7%) | 24 (24.2%) |  |
| **Initial dx to 1st recurrence** |  |  |  |  |  |
| Median (Range) in years | 1.5 (0.8, 5.6) | 1.6 (0.5, 8.8) | 1.7 (0.3, 3.1) | 1.6 (0.3, 8.8) |  |
| <2 years | 29 (78.4%) | 19 (79.2%) | 28 (73.7%) | 76 (76.8%) | 0.8463<sup>1</sup> |
| 2+ years | 8 (21.6%) | 5 (20.8%) | 10 (26.3%) | 23 (23.2%) |  |

<sup>1</sup>Chi-Square p-value; <sup>2</sup>Kruskal-Wallis p-value;

The current analysis included eligible patients from these three trials with lung-only osteosarcoma recurrence following surgical resection. Patients who were considered by their treating physicians to have been refractory to their initial upfront therapy and those without pathologic confirmation of recurrence were excluded in this analysis. Patients enrolled on AOST1321 who had extra-pulmonary recurrence and patients enrolled on AOST0221 who did not complete surgical resection were excluded (Figure 1). For patients enrolled on more than one trial, only data from first recorded therapeutic enrollment was considered.

**Figure 1:**
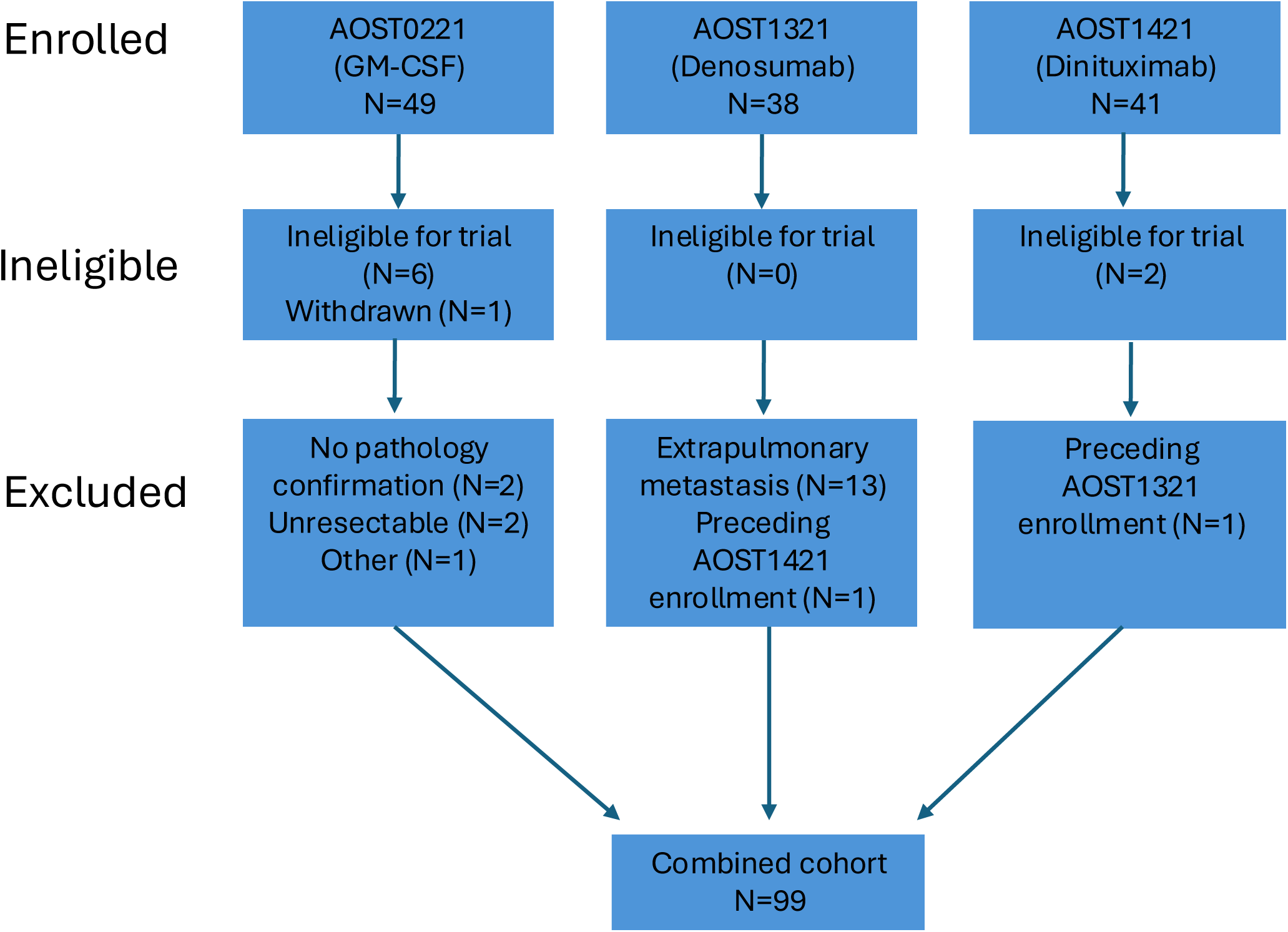
Consort diagram

### Statistical Methods

The primary outcome measure for this study was post-surgery EFS, defined as the date of study enrollment for contemporary trials and the date of last surgical resection for the legacy trial until date of last contact, date of disease progression or detection of disease at a previously uninvolved site or date of death, whichever occurred first. Patients who died or experienced disease progression were considered to have experienced an EFS event; otherwise, the patient was considered censored at the date of last follow-up. Overall survival (OS) was defined analogously as time rendered disease free by surgical resection until death regardless of cause or date of last contact, whichever came first, with censoring at conclusion of OS-time in absence of death. For this report DCS12 was calculated as 12-month EFS.

Potential prognostic factors examined for their influence on risk of an EFS event included study of enrollment, age (years) at enrollment (coded as ≤ 9, 10 to 17, ≥ 18), sex (male *v*. female), race (white, black, other), interval from diagnosis to first recurrence (<2 years *v*. 2+ years), and metastatic disease present at initial diagnosis (yes *v*. no). The probability of remaining event-free (EFS, OS) was estimated by the method of Kaplan and Meier^13^. Tests for equality of survival curves by patient characteristics were conducted using the logrank test^14^. A two-sided P value of 0.05 or less was considered evidence of a significant difference in risk of event across the categories considered.

## RESULTS

### Patients

One hundred and twenty-eight patients were enrolled on AOST0221, AOST1321 and AOST1421. Eight patients (6 from AOST0221 and 2 from AOST1421) were excluded from the analysis due to being ineligible for the trial. An additional six patients from AOST0221 were excluded from this analysis for lack of histologic confirmation of tumor at resection (n=2), patient withdrawal from trial (n=1), unresectable disease prior to protocol specified thoracotomy (n=2), and toxicity from inhaled GMCSF that precluded further protocol specified therapy (n=1). Thirteen additional patients were excluded from AOST1321 because of having extrapulmonary recurrent osteosarcoma at enrollment. Two patients enrolled on more than one trial. Therefore, 99 unique patients across these three studies formed the dataset for this current analysis (Figure 1).

Patient characteristics are listed in Table 1. The majority (58%) of patients were 11-17 years old at enrollment while 36% were 18 years or older. Sixty-one percent of patients were male. Most patients were white, non-Hispanic. The median time from initial diagnosis to first recurrence was 1.6 years (range, 0.3-8.8) and only 24% had metastatic disease at diagnosis. There was no significant difference in these baseline characteristics between the three trials. Numerically, the proportion of males enrolled in the legacy trial (68%) was higher than in the contemporary trials (58% and 55% for AOST1321 and AOST1421, respectively).

### Outcomes

Median EFS for the pooled cohort was 4.83 months [95% Confidence Interval (CI) 3.5-6.3] (Figure 2). EFS at 12 months was 22% (95% CI 15-31%) for all patients included in this analysis. Twelve-month EFS was 14% (95% CI 5-26%) for the legacy trial AOST0221 versus 25% (95% CI 10-43%) and 29% (95% CI 16-44%) for the contemporary trials, AOST1321 and AOST1421, respectively. Comparing the risk of an EFS-event across studies, there was no evidence of difference in risk across the three trials. In this sample, we found evidence to suggest that the risk of an EFS-event is higher for those whose first recurrence occurred within two years of initial diagnosis (Figure 3). While it did not reach statistical significance, male patients may be at increased risk of an EFS-event compared to female patients (Figure 3). We found no evidence of difference in risk of an EFS-event for the following patient related factors in this cohort: age, race, and metastatic status at diagnosis (Supplemental Figure 1).

**Figure 2.**
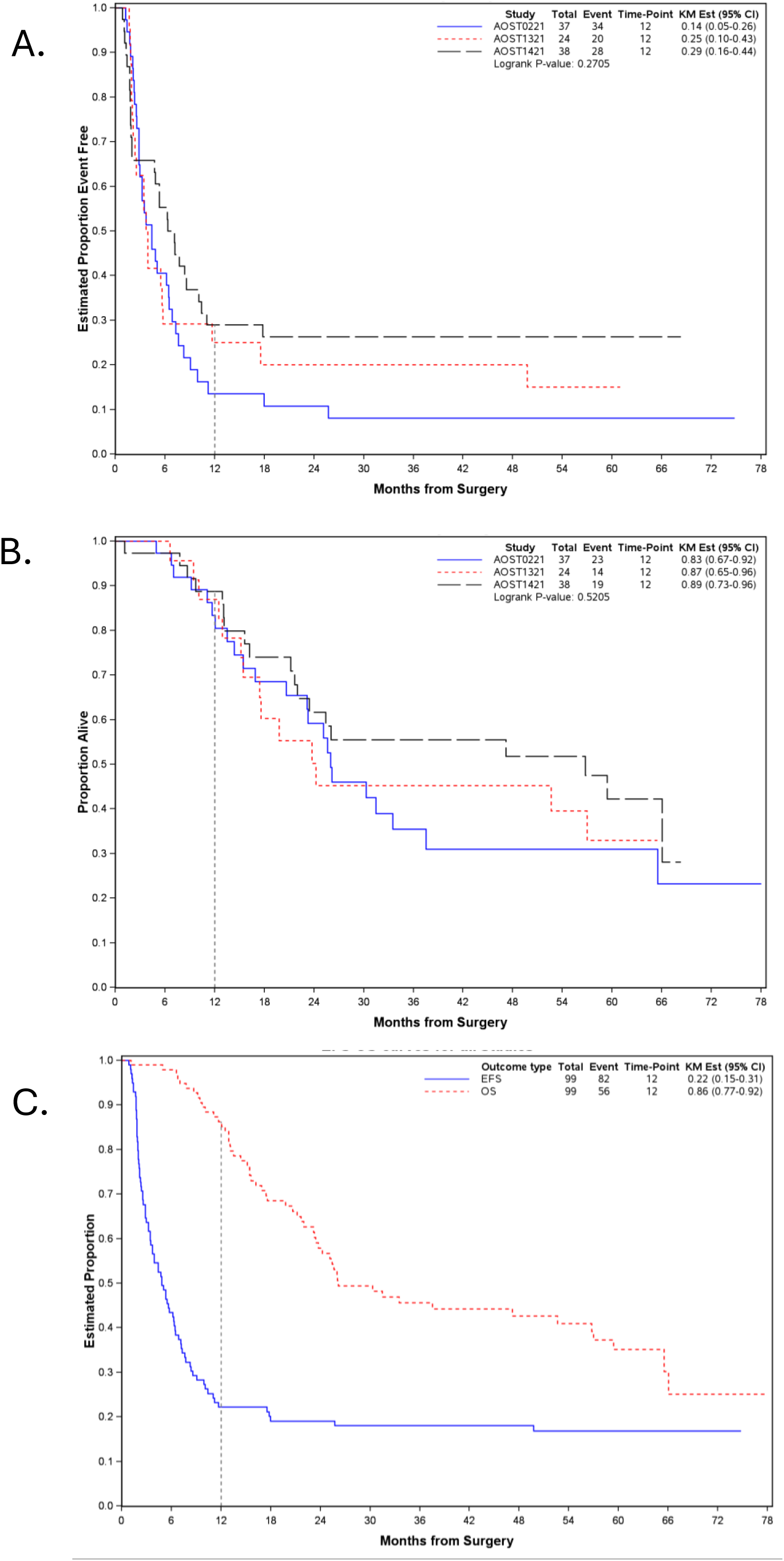
Event Free Survival (A) and overall survival (B) for each of the three trials; EFS and OS for all 99 patients included in the analysis from the 3 trials (C).

**Figure 3.**
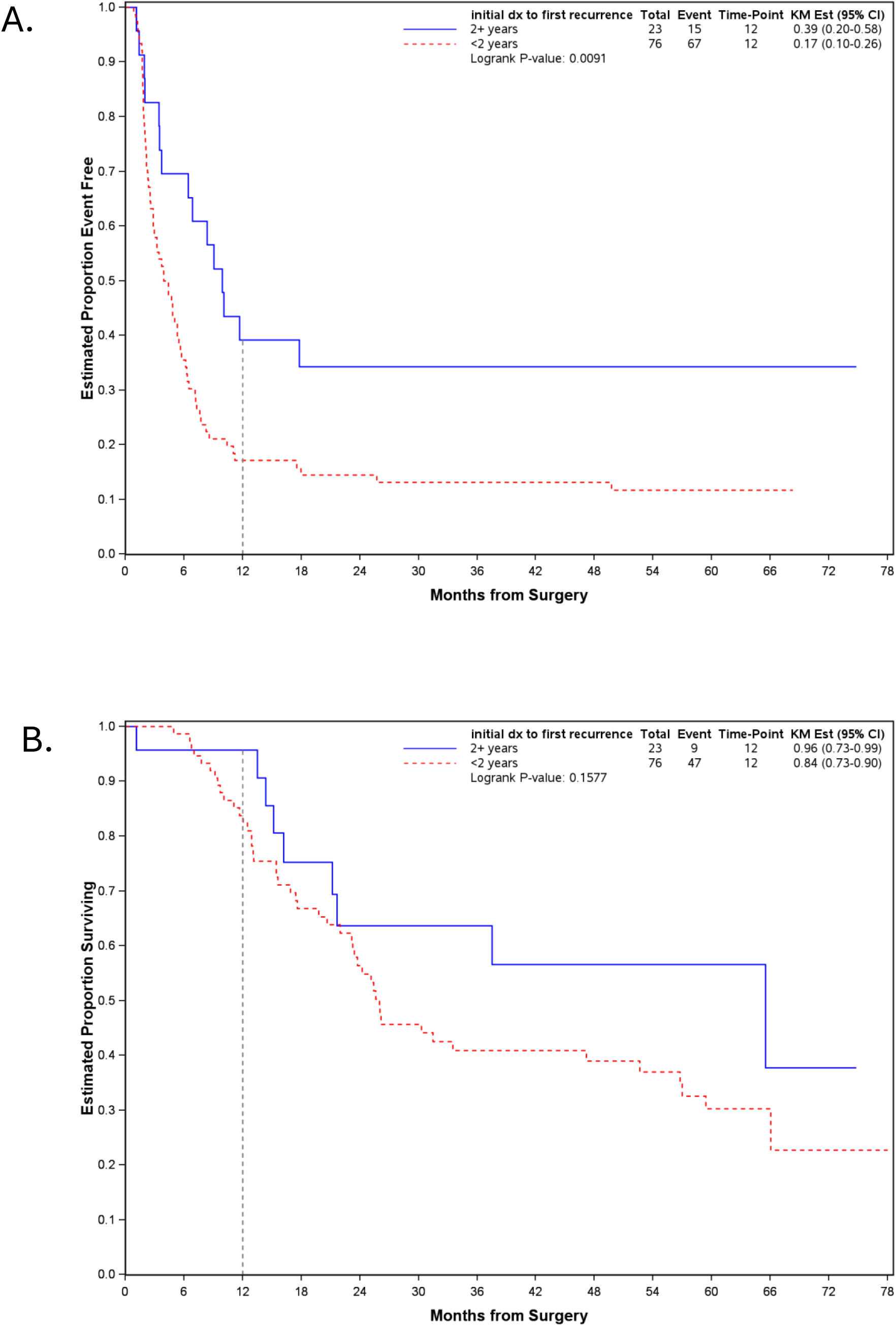
The risk of an EFS event (A) but not death (B) is lower in patients relapsing within 2 years of initial diagnosis.

Median OS for the pooled cohort was 26.2 months (95% CI 23.3-56.8 months). Twelve-month OS was 86% (95% CI 77-92%) for all patients included in this analysis. OS at 12 months was 83% (95% CI 67-92%) for the legacy trial AOST0221 while it was 87% (95% CI 65-96%) and 89% (95% CI 73-96%) for the contemporary trials AOST1321 and AOST1421, respectively. There was no evidence to indicate a statistically significant difference in survival risk across the three trials. The following patient related factors did not statistically affect OS in this cohort: age; sex; race; interval between diagnosis and first recurrence; and metastatic status at diagnosis (Figure 4 and Supplemental Figure 2).

**Figure 4.**
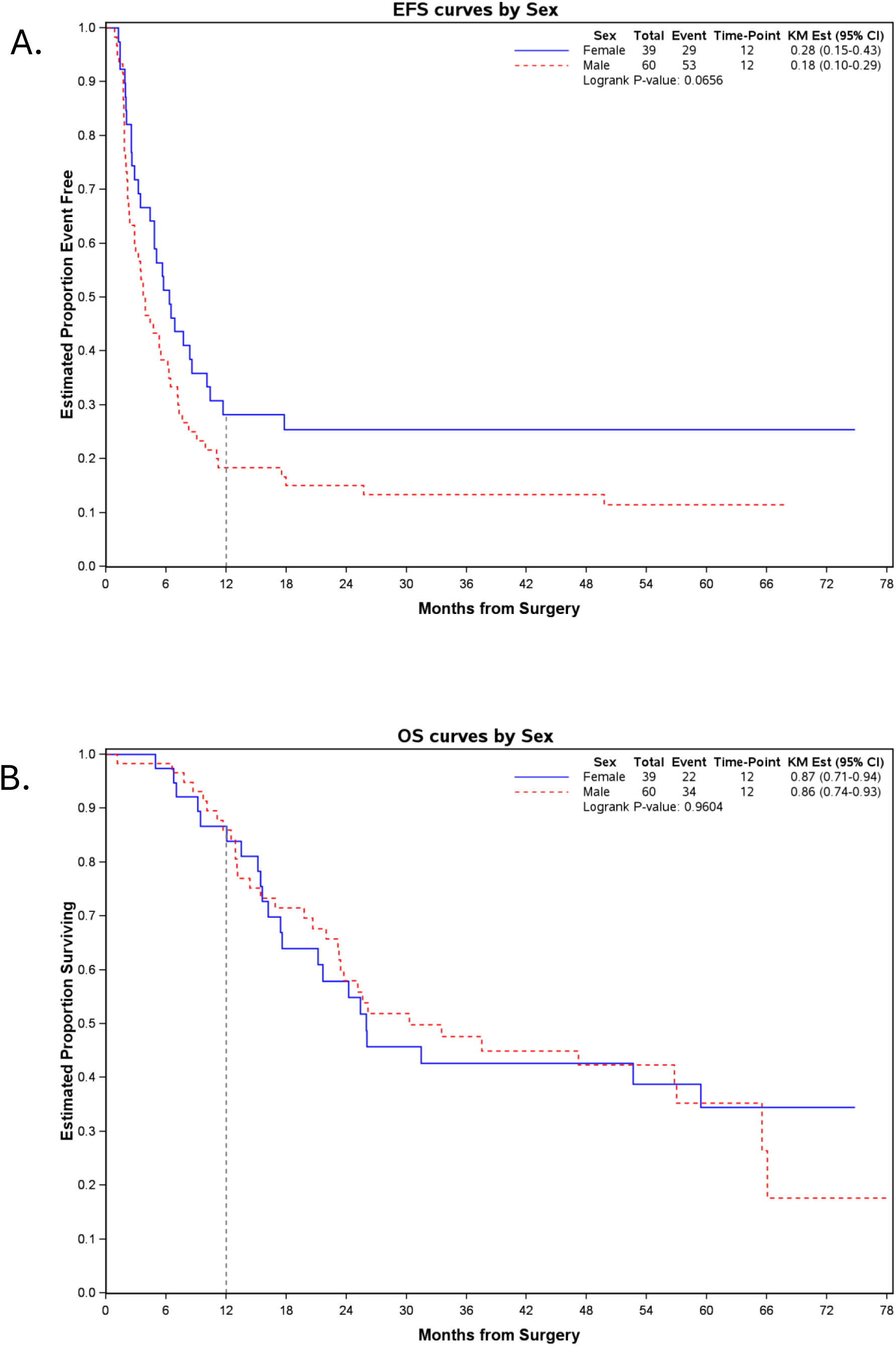
While EFS estimates for male patients are lower than that of female patients, there is no evidence to suggest that the risk of EFS event (A) or death (B) differs by patient sex.

## DISCUSSION

Recurrent osteosarcoma is rare and prognosis remains poor. In such diseases, efficiently run early phase clinical trials are needed to identify promising agents quickly and decrease exposure of patients to agents with little or no evidence of clinical benefit. Well-defined and widely accepted benchmarks can help facilitate the design and conduct of signal finding trials. In our current report, we sought to build on the approach first proposed by Lagmay, et al. In this approach, disease control success at 12 months was proposed as a surrogate endpoint, and a historical benchmark for LOOR-FSR was established.^10^ That approach proved useful, facilitating several signal-finding clinical trials in this patient population over the ensuing decade. As the previously published historical benchmark was based on patients enrolled on a single cooperative group trials between 2004 and 2008, we sought to combine that data with outcomes from two contemporary trials (AOST1321 and AOST1421) to create a more robust benchmark to facilitate the next generation of trials in patients with LOOR-FSR. The analysis of 99 patients enrolled in 3 trials produced a 12-month EFS of 22% with a 95% confidence interval, ranging from 15-33%, that remains wide.

The small patient population, despite combining three trials, and varying (though statistically not different) 12-month EFS in these serial non-randomized phase 2 trials contributes to the wide confidence interval. Possible explanations for the varying 12-month EFS in these serial non-randomized phase 2 trials are several. Advances in supportive care, diagnostic imaging, and surgical techniques may have contributed to the non-significant increase in point estimates of EFS over time. Indeed, the trend in outcome follows chronology, with the lowest 12-month EFS (14%) in the oldest trial and higher 12-month EFS (25 and 29%) in the more recent trials. The true 12-month EFS could be closer to 14% as was seen with AOST0221 and the more recently studied agents (denosumab and dinituximab) have activity with small to moderate effect sizes. Finally, there may be imbalance in the clinical or molecular factors that impact outcome, as we cannot account for such modifiers at this time. For example, we report that time to initial recurrence > 2 years is associated with improved EFS and sex was identified as potentially impacting EFS. Other features such as *MYC* amplification^15,16^ and specific RNA signatures^17^ are associated with poor outcomes after upfront therapy but have not been studied in recurrence. This is the first report to identify a shorter interval from diagnosis to first recurrence as a variable associated with poorer EFS in LOOR-FSR. Prior reports concordant with this finding assessed the correlation in all patients with recurrent osteosarcoma.^5,18^

Despite the failure of the trials reported in this analysis to achieve the primary objective, utilizing this benchmark to determine a time to event endpoint indicative of treatment efficacy is appropriate in certain circumstances. Importantly, this analysis includes only those patients with lung only recurrence who were rendered free of disease with surgery and the updated benchmarks reported herein apply only to this patient population. Thus, any trial utilizing this benchmark should have similar eligibility criteria. Because disease progression is usually detected on imaging rather than by clinical symptoms, trials using this disease control endpoint should have assessment intervals like those in the original trials (Table 1) with disease assessments at 6-8 weeks in the first few cycles. If a single arm design is utilized, it would be essential to plan for a large effect size to achieve sufficient confidence that improvements in outcome were due to the experimental intervention. In fact, the contemporary trials analyzed here were designed in that fashion; the point estimate for efficacy had an odds ratio > 2 compared to the upper 95% CI of the historical benchmark.^13,14^ Unfortunately, no agents have met or exceeded this high efficacy threshold. Overall survival is a more complicated end point in the adjuvant setting as subsequent therapies can impact this outcome measure. Notably, mTKIs were demonstrated to prolong EFS and entered national guidelines in approximately 2020.^6^

There are several scenarios where a randomized design would be a superior approach (to a single arm design) for signal finding trials in patients with LOOR-FSR. If there is concern that requiring a large effect size in this disease context will eliminate agents from further development, then a single-arm design is not appropriate. Second, a single arm trial with a time to event endpoint is less likely to lead to regulatory approval. Finally, clinical trials designed to assess the activity of adding a novel agent to an effective backbone, such as multi-targeted kinase inhibitors (mTKIs) or ifosfamide/etoposide would need to be randomized. This statement agrees with recommendations made by Thebault et al. and van Ewijk et al in their recent discussions of trial designs.^19,20^ The 12-month EFS benchmark presented here could still be informative for randomized trial designs. For example, the benchmark could provide a reference of inefficacy for two randomized arms if the true efficacy of both trial arms is uncertain.

In conclusion, while we were able to update the benchmark with contemporary data and a larger number of patients, there is variability in 12-month EFS and we were unable to confidently identify all factors causing this variability. Given this, single arm designs are unlikely to reliably detect modest treatment effects and are best suited for identifying only large signals of activity. Randomized trials are therefore recommended if the goal is to detect small effect sizes or if combinations with standard relapsed therapies are being studied. A longer interval between diagnosis and first recurrence is associated with improved EFS and male sex may be associated with poorer EFS, highlighting potential stratification factors for future randomized trials or factors required for matching to external controls. An individual-level de-identified dataset containing the variables analyzed in this study can be expected to be available upon request. Requests for access to COG protocol research data should be sent to. Data are available to researchers whose proposed analysis is found by COG to be feasible and of scientific merit and who agree to the terms and conditions of use. Further studies of causes of variability in outcomes in this patient population such as more precise measures of disease burden and biology, are needed.

## Data Availability

An individual-level de-identified dataset containing the variables analyzed in this study can be expected to be available upon request. Requests for access to COG protocol research data should be sent to. Data are available to researchers whose proposed analysis is found by COG to be feasible and of scientific merit and who agree to the terms and conditions of use

## Acknowledgements

We thank the patients and families who participated in the trials analyzed in this manuscript. Research reported in this publication was supported by the National Cancer Institute of the National Institutes of Health under award number U10CA180886 and U10CA180899 to the Children’s Oncology Group. We also thank St. Baldrick’s Foundation. Additional funding from Midnight Riders Pan Mass Challenge team (KAJ)

## Disclaimer

The content is solely the responsibility of the authors and does not necessarily represent the official views of the National Institutes of Health.

## Abbreviations

CI: Confidence Interval
COG: Children’s Oncology Group
CT: Computed Tomography
DCS12: Disease Control Success at 12 months
EFS: Event-Free Survival
GD2: Disialoganglioside2
GM-CSF: Granulocyte Macrophage Colony Stimulating Factor
LOOR-FSR: Lung only osteosarcoma recurrence following surgical resection
NCTN: National Clinical Trials Network
RANKL: Receptor Activator of Nuclear Factor kB Ligand
RECIST: Response Evaluation Criteria in Solid Tumors
OS: Overall Survival

---

**Supplemental Figure 1.**
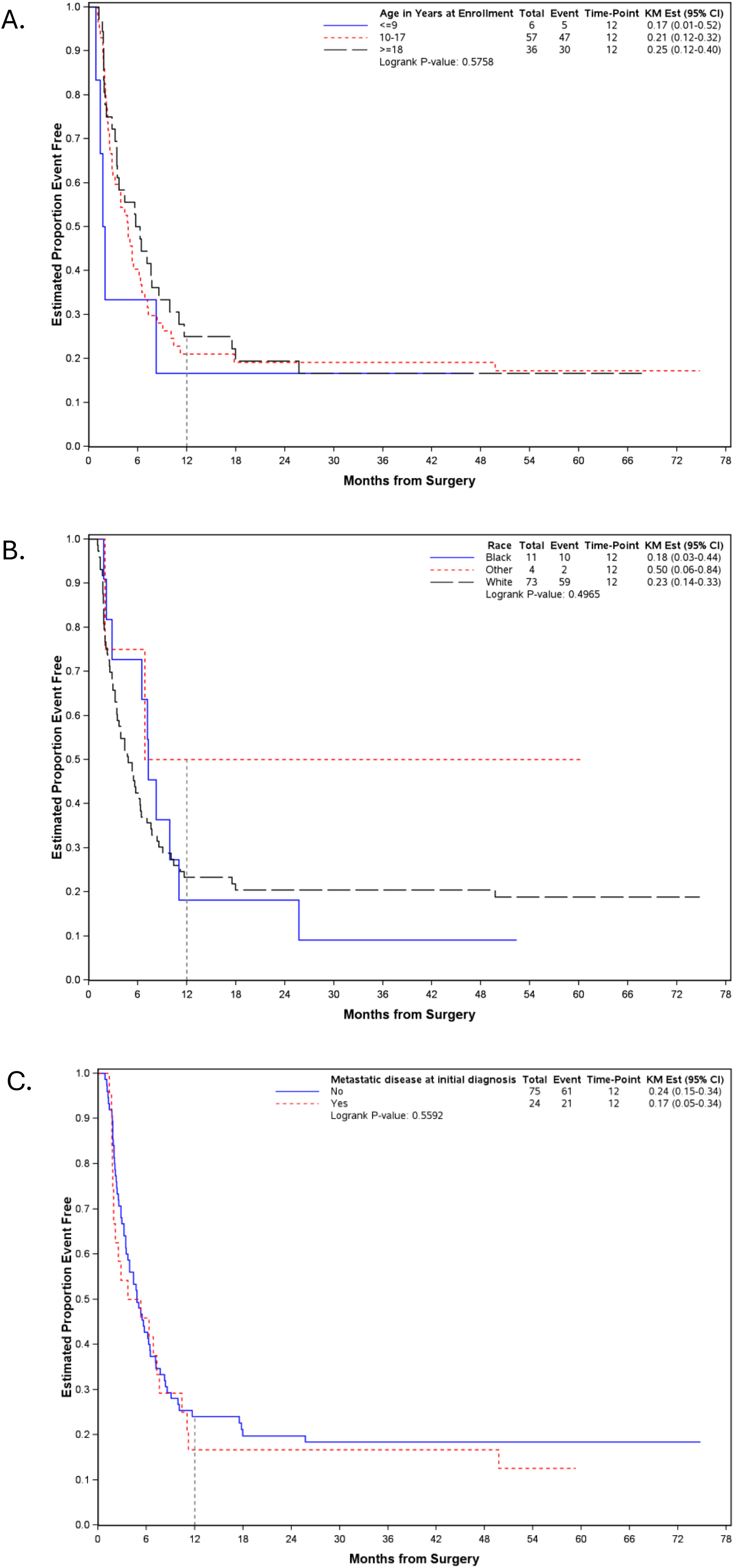
There was no evidence for an association between EFS and age (A), race (B) and metastatic status at diagnosis (C)

**Supplemental Figure 2.**
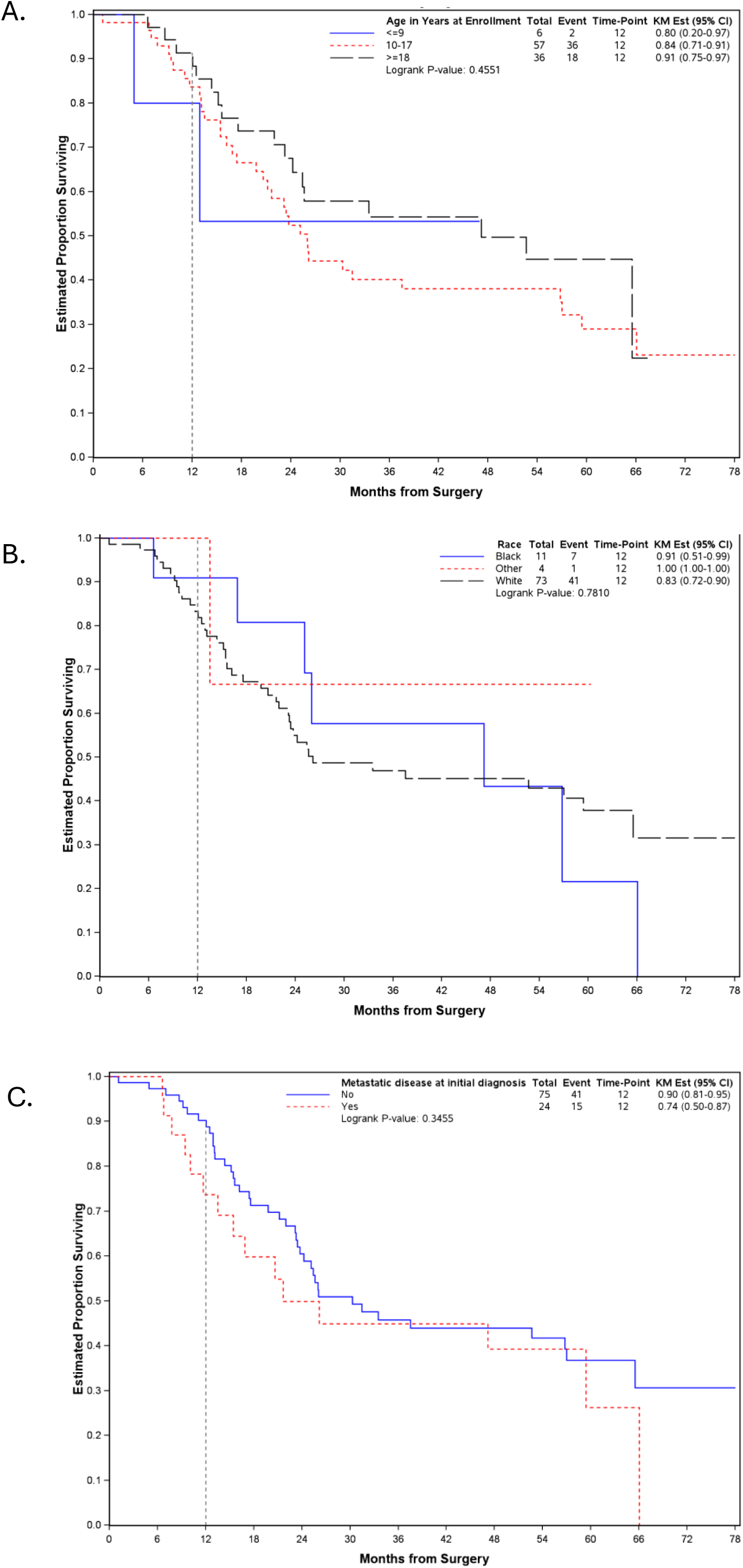
There was no evidence for an association between OS and age (A), race (B) and metastatic status at diagnosis (C)

